# Pandemic Vaccination Strategies: Global Optimization

**DOI:** 10.1101/2023.06.13.23291327

**Authors:** James Thompson, Fernando Benitez-Paez, Omar Guerrero

## Abstract

In the event of a global pandemic, caused by an infectious disease of humans, we consider the scenario in which a safe and effective vaccine is available, with limited supply. We then investigate the optimal distribution of doses across countries and age groups. We employ a system of SIR equations to model the pandemic dynamics and utilize genetic algorithms to identify optimal allocations that minimize either total infections or total deaths. Our findings reveal that strategies aimed at reducing infections differ from those aimed at reducing deaths, with neither strategy advocating proportional dose allocation between countries based on population size. Our findings moreover bring to light potential ethical challenges associated with the implementation of strategy optimization.

## Introduction

Pandemics present destabilizing and deadly threats to human societies, necessitating the development of effective containment strategies to minimize their impact. The ongoing COVID-19 pandemic, for example, has led to hundreds of millions of infections and millions of deaths, with future pandemics being potentially even more devastating. For instance, the highly pathogenic influenza A virus subtype H5N1, an emerging avian influenza virus, has been recognized as a potential pandemic risk. Other coronaviruses besides SARS-CoV-2 could pose similar threats. In this study, we examine a hypothetical pandemic virus causing an infectious disease of humans, characterized by an infectious period and basic reproductive number similar to those of COVID-19. We moreover assume the existence of a safe and effective vaccine, and for the sake of simplicity, we assume that it confers total immunity with complete protection against reinfection. While COVID-19 vaccination rates have varied significantly between countries, due to a combination of supply and demand factors, our model will not consider vaccine hesitancy and instead focus on supply-side limitations.

The study [2] extended a mathematical model of SARS-CoV-2 transmission to identify optimal vaccine allocation strategies within and between countries to maximize averted deaths under limited dose supply. It found that targeting the elderly is optimal under limited supply, while targeting key transmitters provides indirect protection to the vulnerable as supply increases. Assuming a 2 billion global dose supply in 2021, the study reported that allocating doses proportional to population size is close to optimal in averting deaths and that this aligns with ethical principles in pandemic preparedness planning. Using an age-structured model of SARS-CoV-2 dynamics, the study [6] also investigated the global impact of different vaccine sharing protocols, arguing that greater vaccine sharing would have reduced the global burden of disease, assuming that potential increases in infections in previously vaccine-rich countries would be mitigated by maintaining non-pharmaceutical interventions.

In our study, we employ an age-group and country-indexed SIRD model, with cross-country mixing determined by a matrix constructed using international travel data. Given a limited supply of 1 billion doses of a single dose vaccine, policy optimization is performed using differential evolution, a genetic algorithm-based approach.

Our primary finding is that strategic dose sharing between countries, rather than proportional distribution based on population size, can save lives and reduce infections. To minimize deaths, optimal allocation is achieved by sharing the doses according to the size of the vulnerable population in each country. In contrast, minimizing cases necessitates vaccinating regions up to the level of herd immunity sequentially, presenting a moral dilemma as it may require arbitrarily prioritizing one country over another to achieve optimal results.

We conclude that optimization algorithms may offer valuable insights for pandemic preparedness, but their results must be cautiously interpreted and balanced against competing political interests and ethical considerations.

## Methods

In this section, we describe the model and state the underlying assumptions. We provide a mathematical formulation of the model and a link to a public GitHub repository where all the relevant Python code can be found.

### Code

The code is open source and available under a Creative Commons license at the following address: https://github.com/James-Thompson-724/world_sir_model

The code is written in Python, with the optimization algorithm coming from the PyGAD open-source library of genetic algorithms: https://pygad.readthedocs.io/en/latest/

### Model Description

The model is an SIRD model indexed by country and age group. The three age groups appearing in our model are 0-17, 18-64 and 65+. For each country *i* and age group *a*, we denote by *N*_*ia*_ the number of people in age group *a* and country *i*, and by *N*_*i*_ the total population size of country *i*.

Mixing between countries is described by a stochastic matrix *A*_*ij*_. For two countries *i, j*, the entry *A*_*ij*_ denotes the probability that a person from country *i* is mixing inside country *j* on a given day. This matrix is approximately the identity, with the difference representing the impact of international travel. Recall that the matrix *A*_*ij*_ being stochastic means that its rows sum to one. We construct this matrix as a combination of two other stochastic matrices *B*_*ij*_ and *C*_*ij*_. The matrix *B*_*ij*_ corresponds to air travel while the matrix *C*_*ij*_ corresponds to travel by any means other than air travel. We use air travel data to estimate *B*_*ij*_ directly, while to determine *C*_*ij*_ we impose a theoretical model. In particular, we suppose that each day a proportion *c* of the population of each country travels to neighbouring countries, with the number of travellers *cN*_*i*_ being distributed among the neighbouring countries according to their population sizes. In particular, denoting by *a*_*ij*_ the adjacency matrix equal to 1 for neighbouring regions and 0 otherwise, we suppose that

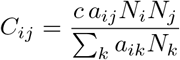

and finally obtain the matrix *A*_*ij*_ using the formula

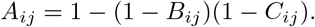

To describe mixing between age groups, for each country *i* and age groups *a* and *b*, we denote by *M*_*iab*_ the corresponding daily mixing probability. For simplicity, we assume that

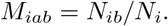

Our SIRD model records changes to the quantities *S*_*ia*_(*t*), *I*_*ia*_(*t*), *R*_*ia*_(*t*) and *D*_*ia*_(*t*), representing the numbers of people who are susceptible, infected, removed and dead, respectively, in region *i* in age group *a* at time *t*, where by removed we mean either recovered or vaccinated. We denote by *β* and *γ* the transmission and recovery rates, respectively, and for each age group *a*, we specify an infection fatality ratio *µ*_*a*_. We will assume *γ*^−1^ = 9 days with *β* = 0.2723 days^−1^, so that in the absence of international travel and age structure the model reduces to a standard SIRD model with *R*_0_ = 2.45, as in [7]. The dynamics of the system are then determined by the following set of ordinary differential equations, together with appropriate initial conditions, for each country *i* and age group *a*:

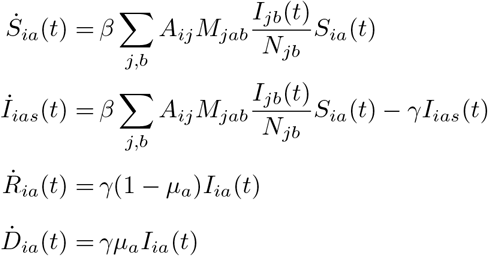

for *t >* 0. Note that in the case of only one country and only one age group we have *A*_*ij*_ = 1 and *M*_*jab*_ = 1 and these equations reduce to those of a standard SIRD model. To represent the impact of vaccination, a number is subtracted from the susceptible compartment and added to the removed compartment each day. We solve the equations numerically, using a forward Euler scheme, over a time interval of length 365 days, and having done so, we record as the total cost either the total number of cases or the total number of deaths. For a given supply of vaccine, the problem is then to find the allocation of doses, to each country and to each age group within each country, that minimizes either total cases or total deaths.

### Optimization

We employ genetic algorithms to identify optimal strategies in our study. Genetic algorithms begin with a randomly generated set of candidate solutions and use biologically inspired operations like mutation, crossover, and selection to evolve the solution population over multiple generations, ultimately achieving higher fitness. In our case, a solution determines the number of people vaccinated in each country and age group, while a solution’s fitness is determined by the reduction in deaths or cases compared to the baseline scenario in which there is no vaccination. The search continues until terminal conditions are met, which, in our case, happens after 10,000 generations, allowing the fitness curve to stabilize, indicating that the solutions are approximately optimal.

It is important to note that these algorithms are stochastic and yield only approximately optimal solutions. Moreover these solutions may only be local optima and may not be unique. Solutions should therefore be interpreted with these limitations in mind. Nevertheless, genetic algorithms are powerful tools that provide valuable insights into numerous problems, including optimal pandemic strategies, as demonstrated in our study.

### Input Data

Data on population size and age structure are sourced from [8]. Air travel is configured using the model presented in [3], which features an open-access passenger flow matrix for the global air network in 2010, later refined in [4] to provide monthly estimates. Age-dependent infection-fatality ratios are taken to be those for COVID-19 appearing in [1]. For time-dependent supply scenarios, we assume that each country can vaccinate at most 0.6% of its population each day, this percentage being based on global vaccinate rates during the height of the COVID-19 pandemic [5].

## Results

Before optimizing the global model, let us begin with a simple example. In particular, suppose we have only two countries, Country A and Country B, and suppose for simplicity that there is no mixing between these countries. For the values of *β* and *γ* given in the previous section, the herd immunity threshold for each country is then *p*_*c*_ := 1 − 1*/R*_0_ = 0.59. Let us suppose that Country A and Country B both have populations of size *N* = 1, 000, 000, and that epidemics begin simultaneously in each country with one initial case. Suppose these countries have a shared stockpile of *d* doses of a vaccine, and that the delivery system in each country allows for instantaneous distribution on day zero. If *d <* 2*N*, meaning that there are insufficient doses for everyone, how should the doses be divided to minimize total cases?

The answer depends on *d*. If 2*p*_*c*_*N* ≤ *d*, then there are enough doses to achieve herd immunity in both countries, and the optimal strategy is to share the doses equally. However, if *p*_*c*_*N* ≤ *d <* 2*p*_*c*_*N*, only one country can achieve herd immunity, and the optimal strategy is to give *p*_*c*_ ∗ *N* doses to one country with the remainder going to the other country. If 0 ≤ *d < p*_*c*_*N*, the optimal strategy is to give all the doses to only one of the two countries. These three cases are illustrated below in Figure 1.

**Figure 1.**
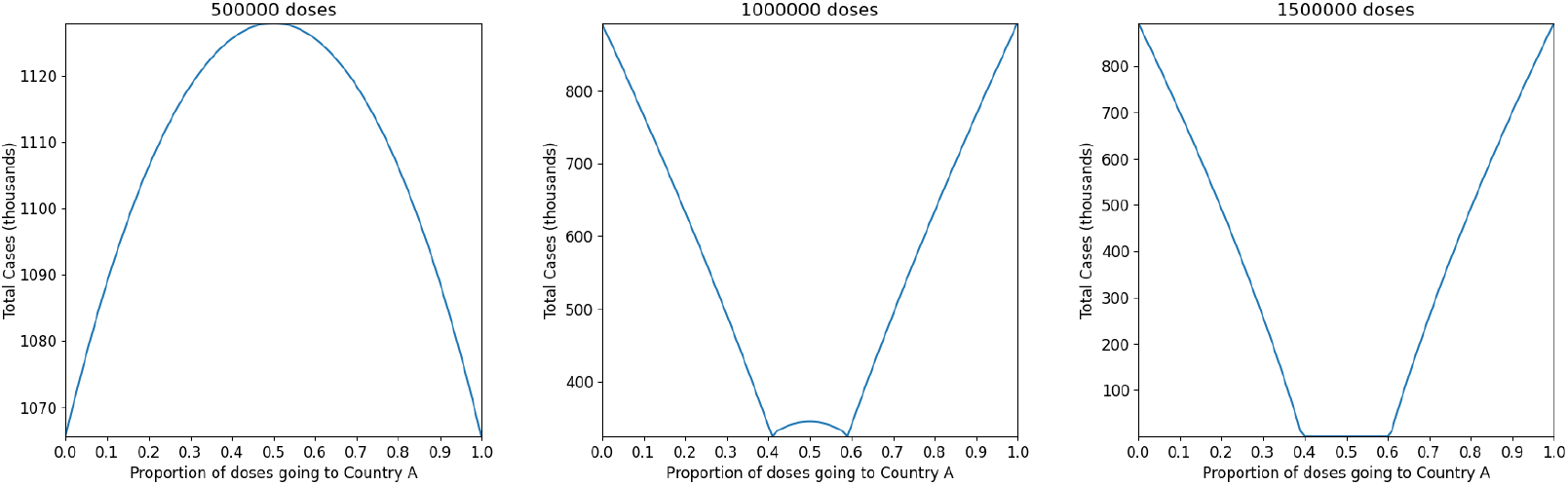
On the left, with a small stockpile of doses, cases are minimizes by giving all the doses to only one of the two countries. In the center, for a medium sized stockpile, cases are minimized when one country receives enough doses to achieve herd immunity, with the remainder going to the other country. For a large stockpile, both populations can be fully protected and an equal sharing of the doses achieves this.

We see from this example that for stockpiles smaller that 2*p*_*c*_*N*, there is a moral dilemma, since in this case sharing the doses equally is sub-optimal, with the optimal allocation instead requiring the arbitrary selection of one country over the other. This is because the impact of vaccination depends non-linearly on the proportion of the population vaccinated. Extending this example to the case where Countries A and B have different population sizes, we find that with a limited supply of doses, the optimal allocation often favours the smaller of the two countries, due to it being easier to achieve herd immunity there.

In this example we minimized only cases, as opposed to deaths. As shown in [2] for the COVID-19 vaccines, the optimal strategy for minimizing deaths with limited supply involves prioritizing the most vulnerable individuals, but that targeting transmission provides indirect protection to the vulnerable as supply increases. Therefore, even when minimizing deaths, the herd immunity dilemma outlined in this example will become relevant as supply increases.

We now turn to the global model, and suppose the pandemic begins with 10,000 initial cases in China and that a stockpile of 1 billion doses of vaccine is to be allocated on day zero between countries. We first run the genetic algorithm in such a way that minimizes cases. We find that the solution produced by the genetic algorithm prioritizes countries with small populations, in which it is relatively easy to achieve herd immunity, as predicted by the example. The solution does not, in particular, share the 1 billion doses equally between countries. Note, however, that subdividing countries countries in the model will result in different solutions.

We now run the genetic algorithm in such a way that it minimizes deaths. For each country, age group infection fatality ratios are calculated using the COVID-19 age-dependent infection fatality ratios reported in [1], adjusted to the age structure of each country. We find that the optimal solution vaccinates the elderly first. The solution therefore allocates a disproportionately large share of the doses to high-income countries in Europe, North America and East Asia, since these countries have the oldest populations. Were the infection fatality ratio to be highest among children, as opposed to the elderly, then the optimal solution would allocate a disproportionately large share to countries with the youngest populations, many of which are low and middle-income countries in Africa. The herd immunity effect is secondary with such a limited stockpile, so once the elderly have all been vaccinated, the remaining doses go to small countries, where herd immunity is most easily achieved.

Moreover, even if a safe and effective vaccine is available, as we have assumed, significant numbers of people around the world will in reality be either unable or unwilling to be vaccinated. Factoring in such limits on supply and demand, with the supply of doses being distributed over time rather than all available on day zero, and using data on COVID-19 vaccine coverage up to April 2022 to model limits on coverage, the optimization of our model results in solutions exhibiting even greater disparities between countries, particularly when minimizing deaths.

In scenarios with extremely low daily vaccine supplies, the international travel network gains increasing importance. We find that in this setting the optimal solution tends to favor countries situated furthest from the pandemic’s origin, as they have more time before their epidemic peaks compared to countries closer to the origin. This extended time frame makes it easier to achieve herd immunity in these distant countries before the peak occurs.

The full version of our model supports multiple strains and non-pharmaceutical interventions, including border closures and lockdowns. These interventions apply a selection pressure on the strains, leading to a complex interplay of dynamics. Simultaneously optimizing these interventions and vaccination strategies can result in even more disparate outcomes than those remarked upon above.

## Discussion

In this study, we considered the situation in which a stockpile of 1 billion doses of a safe and effective vaccine is available for use against a pandemic virus. Using genetic algorithms, we optimized the global distribution of these doses between countries and between age groups within countries. The optimization was first performed in such a way as to minimize total cases, before then being performed in such a way as to minimize total deaths.

Our results imply that when minimizing cases, the optimal strategy involves vaccinating countries or geographical regions one at a time, each to the level of herd immunity, starting with the region in which this target can be most easily achieved. Regions vaccinated to this level could then be free to interact with one another, in the absence of any social restrictions, while the vaccination program continues to work through the remaining regions as far as it can. To minimize deaths, the optimal strategy involves vaccinating the highest risk groups first, after which remaining doses are allocated in such a way that maximizes herd immunity. Since the proportion of high risk individuals varies between countries, neither strategy involves sharing the doses proportionally by total population size.

We have therefore shown that systematic optimization of policy in a pandemic context can lead to solutions that may conflict with ethical principles. Minimizing deaths, or other measures, using algorithms may seem appealing, but could result in questionable solutions to moral dilemmas similar to the trolley problem. More generally, the application of artificial intelligence to social problems requires great care. Depending on the choice of optimizing function, the solution might involve ruthlessly discriminating one group or region over another. On the other hand, the solutions obtained by such methods can offer valuable insights, the logic of which can be integrated into an effective policy.

Furthermore, care is needed when employing heuristic methods, such as genetic algorithms, that yield only approximate solutions. The optima may only be local and may not be unique. For instance, the solution might contain arbitrary random choices.

The pandemic model presented in this study is a relatively simple equation-based model. In contrast, the individual-based model, *Pandemia*, is a more sophisticated global pandemic model that incorporates international travel, within-country spatial dynamics based on a modified gravity model, seasonality, multiple strains, waning and cross immunity, multiple vaccines with varying efficacy, as well as a broad range of interventions. The code for this model can be found at: https://github.com/PandemiaProject/pandemia

Although this individual-based model is more computationally demanding, it remains within the scope of machine learning methods such as genetic algorithms or reinforcement learning. Currently, work is in progress to extend the optimization methods applied in this paper, from the equation-based model outlined above to the individual-based model, Pandemia.

## Data Availability

All data produced are available online at:
https://github.com/James-Thompson-724/world_sir_model

## References

[1] Variation in the COVID-19 infection-fatality ratio by age, time, and geography during the pre-vaccine era: a systematic analysis. The Lancet, 399(10334):1469–1488, 4 2022.

[2] Alexandra B. Hogan et al. Within-country age-based prioritisation, global allocation, and public health impact of a vaccine against SARS-COV-2: A mathematical modelling analysis. Vaccine, 39(22):2995–3006, 2021.

[3] Zhuojie Huang, Xiao Wu, Andres J. Garcia, Timothy J. Fik, and Andrew J. Tatem. An open-access modeled passenger flow matrix for the global air network in 2010. PLOS ONE, 8(5):1–9, 05 2013.

[4] Liang Mao, Xiao Wu, Zhuojie Huang, and Andrew J. Tatem. Modeling monthly flows of global air travel passengers: An open-access data resource. Journal of Transport Geography, 48:52–60, 2015.

[5] E. Mathieu, H. Ritchie, E. Ortiz-Ospina, et al. A global database of COVID-19 vaccinations., 2021.

[6] Sam Moore, Edward M. Hill, Louise Dyson, Michael J. Tildesley, and Matt J. Keeling. Retrospectively modeling the effects of increased global vaccine sharing on the COVID-19 pandemic. Nature Medicine, 28(11):2416–2423, 11 2022.

[7] James Thompson and Stephen Wattam. Estimating the impact of interventions against covid-19: From lockdown to vaccination. PLOS ONE, 16(12):1–51, 12 2021.

[8] Department of Economic United Nations and Population Division Social Affairs. World population prospects 2019, online edition. rev. 1., 2019.

